# Cross-Sectional Evaluation of the Alzheimer’s Disease and Related Dementia (ADRD) Risk-Reducing Benefits Associated with HMG-CoA Reductase Inhibitors, or Statins

**DOI:** 10.1101/2025.04.03.25325202

**Authors:** Mohin Chanpura, Zeba M. Khan, Ayse Akincigil

**Affiliations:** Center for Health Outcomes, Policy, and Economics (HOPE) Rutgers, The State University of New Jersey 160 Frelinghuysen Rd., Piscataway, NJ 08854; School of Social Work 120 Albany St., New Brunswick, NJ 08901; Institute for Health, Health Care Policy, and Aging Research Rutgers, The State University of New Jersey 112 Patterson St., New Brunswick, NJ 08901

## Abstract

**BACKGROUND:** HMG-CoA reductase inhibitors (statins) are well-established as an effective pharmacotherapy for dyslipidemia, a condition thought to trigger excess amyloid beta (Aβ) aggregation. Statins may therefore also offer prophylaxis against Alzheimer’s Disease and Related Dementias (ADRD).

**METHODS:** To assess associations between statin use and ADRD risk, we conducted a secondary analysis of United States Medicare Current Beneficiary Survey (MCBS) data from 2019-2020. Each respondent with ADRD was matched with two other persons based on key demographic characteristics to create cases and comparisons, making for a total of 3,468 Medicare beneficiaries included in the analysis. Beneficiary ADRD diagnosis statuses were identified using MCBS’s self-reported Health Status and Functioning Questionnaire and/or the Health Status section of the MCBS Facility Instrument. Prescription utilization data, as published in the MCBS PME module, was used to identify statin users and nonusers. Associations between statin use and ADRD diagnosis were assessed via Fisher’s exact tests.

**FINDINGS:** Statin utilization was associated with significantly reduced ADRD risk (OR 0.68; p<0.0001). Rosuvastatin was associated with significantly reduced ADRD risk relative to atorvastatin (OR 0.66; p=0.0193). Additionally, hydrophilic statins were associated with significantly reduced ADRD risk relative to lipophilic statins (OR 0.77; p=0.0394).

**CONCLUSIONS:** While statin use was found to correlate to significantly reduced ADRD risk, longitudinal research remains necessary to confirm that statins are indeed effective prophylactics against ADRD.

## INTRODUCTION

Alzheimer’s disease and related dementias (ADRD) comprise a class of neurodegenerative conditions that initially manifest as short-term memory loss before evolving into disorientation, mood swings, depression, self-neglect, loss of bodily functions, and ultimately death.^1^ While the causal pathway of this class of dementias is currently disputed, the leading hypothesis states that ADRD presents once a critical mass of amyloid beta (Aβ) has accumulated extracellularly within the brain, disrupting neuronal functioning and connectivity.^2,3^ As Aβ peptides derive from the amyloid precursor protein (APP) through a process regulated by cholesterol transport, ADRD is popularly thought to be a byproduct of dyslipidemia.^4^ Singh-Manoux et al. lent further credence to this theory in 2008 when, utilizing data from Britain’s Whitehall II study, it found low HDL-C levels (<40 mg/dL) were significantly associated with declining short-term memory in participants ages 55-61.^5^

Until 2021, approved treatment options for mild-to-moderate Alzheimer’s disease in the United States had been limited to galantamine, rivastigmine, and donepezil: acetylcholinesterase inhibitors shown to have little efficacy in improving cognitive symptoms associated with ADRD.^6^ In June 2021, the U.S. Food and Drug Administration controversially approved Biogen’s aducanumab for use in Alzheimer’s patients with mild cognitive impairment (MCI).^7^ A first-in-class monoclonal antibody targeting aggregated Aβ, aducanumab was panned by regulatory experts as showing insufficient evidence to demonstrate clear benefits to patients.^8,9^ The regulatory backlash also extended to other Aβ-targeting monoclonal antibodies, including Biogen/Eisai’s lecanemab, which was also controversially approved by the FDA in January 2023 for ADRD-associated MCI, and Eli Lilly’s donanemab, for which the FDA issued a Complete Response Letter in 2023 requesting clinical data from ≥100 patients on at least 12 months of continuous therapy before eventual approval in July 2024.^10–12^ Additionally, with Biogen/Eisai having recently set wholesale acquisition costs (WACs) for aducanumab and lecanemab at $28,200 and $26,500 per-patient-per-year, respectively, and Lilly now estimating donanemab’s 12-month course of therapy cost to be $32,000 per-patient, regulators have also taken issue with the seemingly exorbitant pricing schemes attached to these antibodies.^12–14^ The Centers for Medicare & Medicaid Services, for example, responsible for administering health insurance to nearly all ADRD patients in the United States, declared in 2022 that “there is not currently enough evidence of [Aβ-targeting mAbs] demonstrating improved health outcomes to say that [treatment] is reasonable and necessary for people with Medicare,” and have since remained committed to this position regarding the class’s coverage.^15^

In contrast to Aβ-targeting mAbs, HMG-CoA reductase inhibitors, commonly referred to as “statins,” collectively comprise a well-established, low-cost class of effective treatments for dyslipidemia, with generic versions of atorvastatin, pravastatin, rosuvastatin, and simvastatin all available to pharmacies in the United States for yearly WACs below $20/patient as of January 2024.^16–20^ As HMG-CoA reductase plays a key role in the biosynthesis of cholesterol, statins are also believed to prevent cerebral Aβ aggregation through their inhibitory effect on cholesterol production.^21,22^ While several meta-analyses have examined the effects of statin use on the development of Alzheimer’s disease over the last decade, few studies published within this timeframe have utilized novel data to investigate this relationship. One such study, published in 2020 by Barthold et al., analyzed data collected from nearly 694,000 Medicare beneficiaries between 2007 and 2014 and found that statins, in combination with certain hypertensives, were indeed correlated with significantly reduced ADRD risk. Barthold et al.’s paper was also significant in that it was the first and only publication to date to make statistical comparisons between the varying degrees of ADRD risk reduction associated with individual statin drugs.^23^

The primary objective of this study was to address the dearth of recent literature on statins and Alzheimer’s risk by retrospectively analyzing recent data from CMS’s Medicare Current Beneficiary Survey (MCBS) to determine whether statins as an overall drug class were associated with significantly reduced ADRD risk among older adult users versus nonusers. We additionally compared the ADRD risk reductions associated with individual statin drugs to discern whether certain statins correlated to significantly less ADRD risk than others.

## METHODS

### Study design

This (sub-)study was a retrospective, observational, matched case-control analysis of anonymized Medicare Current Beneficiary Survey data from the years 2019 and 2020. The parent study, titled “Chronic Illness Care for Medicare Beneficiaries” (Study ID: Pro2019001406), was approved by the Rutgers Institutional Review Board with the aim of examining patterns in diagnosis, treatment, and costs of chronic illnesses among Medicare beneficiaries, as well as associated outcomes. MCBS data was accessed numerous times between 02/01/2023 and 02/28/2023 for feasibility analysis, and again between 05/23/2023 and 07/14/2023 for case/control identification, matching, and statistical analysis.

### Data source

The Medicare Current Beneficiary Survey is a longitudinal, multipurpose panel survey of a nationally representative sample of Medicare beneficiaries administered by the Centers for Medicare & Medicaid Services.^25^ Each year, the MCBS surveys a cohort of approximately 15,000 beneficiaries, with over 1 million interviews conducted to date since the survey’s inception.^25^ Distinguishing features of the MCBS include triangulation of prescription utilization data from both self-reported survey responses and administrative claims, complete source of payment information, a rotating panel design, the inclusion of beneficiaries living in long-term care facilities, oversampling of special populations, the inclusion of Medicare Advantage beneficiary data, and rapid data collection for emerging needs such as information on the impact of COVID-19.^27^ Complete annual MCBS data sets, excluding specific direct identifiers as defined by the HIPAA Privacy Rule, can be requested by researchers through filing a limited data set data use agreement (DUA) with CMS.

### Study population

The study population was limited to 2019 and 2020 MCBS respondents at least 45 years of age with: 1) adequate responses to the Demographics and Income Questionnaire, 2) either self- or facility-reported health status inventories, and 3) corresponding prescription utilization data available in MCBS’s Prescribed Medicine Event (PME) module.

### Measures

The study assessed three categories of measurement: ADRD diagnosis status, statin utilization, and demographic characteristics. Diagnosis status was identified through either self-reported survey questions asking beneficiaries if they’d ever been told by a doctor or other health professional that they had Alzheimer’s disease or dementia, or facility reported variables asking responders if the studied beneficiaries had either conditions checked off as active diseases on their minimum data set (MDS) assessment.^28^ The minimum data set is a standardized assessment created by CMS to aid in the facilitation of care management in nursing homes and non-critical access hospital bed swings.^29^

The following demographic characteristics were used to match all identified persons with ADRD with control beneficiaries (those without self or facility reported ADRD diagnoses) at a 1:2 ratio: sex, race (and, if possible, ethnicity), age stratum, and income level. Ethnicities used to match cases with controls (if a case patient responded as identifying as Asian, Hispanic, or Native Hawaiian/Pacific Islander) included Asian Indian, Chinese, Filipino, Japanese, Korean, Vietnamese, Mexican, Puerto Rican, Cuban, and Native Hawaiian.^28^ Beneficiaries were matched using the following age strata: 45-64; 65-69; 70-74; 75-79; 80-84; and 85+, and by the following income levels: $0-4,999; $5,000-9,999; $10,000-14,999; $15,000-19,999; $20,000-24,999; $25,000-29,999; $30,000-39,999; $40,000-49,999; $50,000-59,999; $60,000-79,999; $80,000-99,999; $100,000-$119,999; $120,000-$139,999; and $140,000+.^28,30^ This was not a probabilistic match; we conducted exact matching when possible. If less than two respondents were found to match a case across all 4-5 key demographic characteristics of interest, controls were first identified in adjacent or closest income levels, and subsequently in adjacent or closest age strata, or, if applicable, different ethnicities within the same race. If more than two matches were identified across all key demographic characteristics, a random number generator was used to select the controls. Once all selected cases and controls had been identified, chi-squared tests were utilized to test whether the control group was equivalent to the case cohort in terms of age, income, and ethnicity, as well as education and hypertension prevalence.

Finally, prescription utilization data, as published in the PME module, was used to distinguish statin users from nonusers. For the purposes of this study, “statin user” was defined as any MCBS respondent who reported taking one or more of the following medications: atorvastatin (including amlodipine/atorvastatin), fluvastatin, lovastatin, pitavastatin, pravastatin, rosuvastatin, or simvastatin (including ezetimibe/simvastatin).

### Statistical analyses

Once all selected beneficiaries were designated either statin users or nonusers, the data was organized into multiple 2x2 contingency tables to calculate odds ratios of Alzheimer’s, Other Dementia, and ADRD (Alzheimer’s + Other Dementia) diagnosis among statin users versus nonusers. Fisher’s exact tests were simultaneously employed to determine the significances of the derived odds ratios, and thereby whether statins as an overall drug class were associated with significantly reduced ADRD risk in treated respondents. Next, in order to compare ADRD risk reductions associated with individual statin drugs, all identified statin users were further categorized based on their prescribed statin (i.e., atorvastatin users, pravastatin users, etc.), and the data were again organized into multiple 2x2 contingency tables first comparing users of each particular statin drug to all statin nonusers, then those of one statin drug to those of another, and finally users of hydrophilic statins to those of lipophilic statins. Fisher’s exact testing was again simultaneously employed in each analysis to determine the significances of the associated odds ratios. Due to an insignificant sample size of users (n=92 in total), fluvastatin, lovastatin, and pitavastatin were excluded from the first two sub-analyses of individual statin drug effects. All statistical analyses within the study were performed at the 0.05 significance level using SAS® version 9.4.

## RESULTS

In all, 1,684 unique persons living with Alzheimer’s and/or Other Dementia were identified from MCBS’s 2019-2020 data, 27 of which were immediately excluded based on age (ranging from 24 to 44). Out of the remaining 1,657 cases, 501 were subsequently also excluded from the analysis due to missing corresponding data on prescription drug utilization, yielding a total of 1,156 eligible cases included in the analysis.

**Table 1** displays a crosstabulation of all cases and controls included in the analysis, broken down by statin use versus nonuse. In total, 1,898 out of the 3,468 included cases and controls were identified as statin users. **Tables 2** and **3** display similar crosstabulations of all controls and all cases diagnosed with Alzheimer’s disease, and all controls and all cases diagnosed with other forms of dementia, respectively, broken down by statin use. Overall, statin use was found to correlate to significantly reduced ADRD risk, with odds of diagnosis with Alzheimer’s disease reduced by 36.6% (OR 0.63, 95% CI: 0.52-0.78; p<0.0001), and odds of diagnosis with other forms of dementia reduced by 29.5% (OR 0.71, 95% CI: 0.60-0.83; p<0.0001).

**Table 1:** Diagnosis of ADRD (Alzheimer’s + Other Dementia) among all statin users versus nonusers.

|  | Diagnosed ADRD | No Diagnosis | Total |
| --- | --- | --- | --- |
| Statin User | 558 (29.4%) | 1340 (70.6%) | 1898 |
| Statin Nonuser | 598 (38.1%) | 972 (61.9%) | 1570 |
| Total | 1156 | 2312 | 3468 |

OR: 0.68 (0.59, 0.78); $p < 0.0001$

**Table 2:**
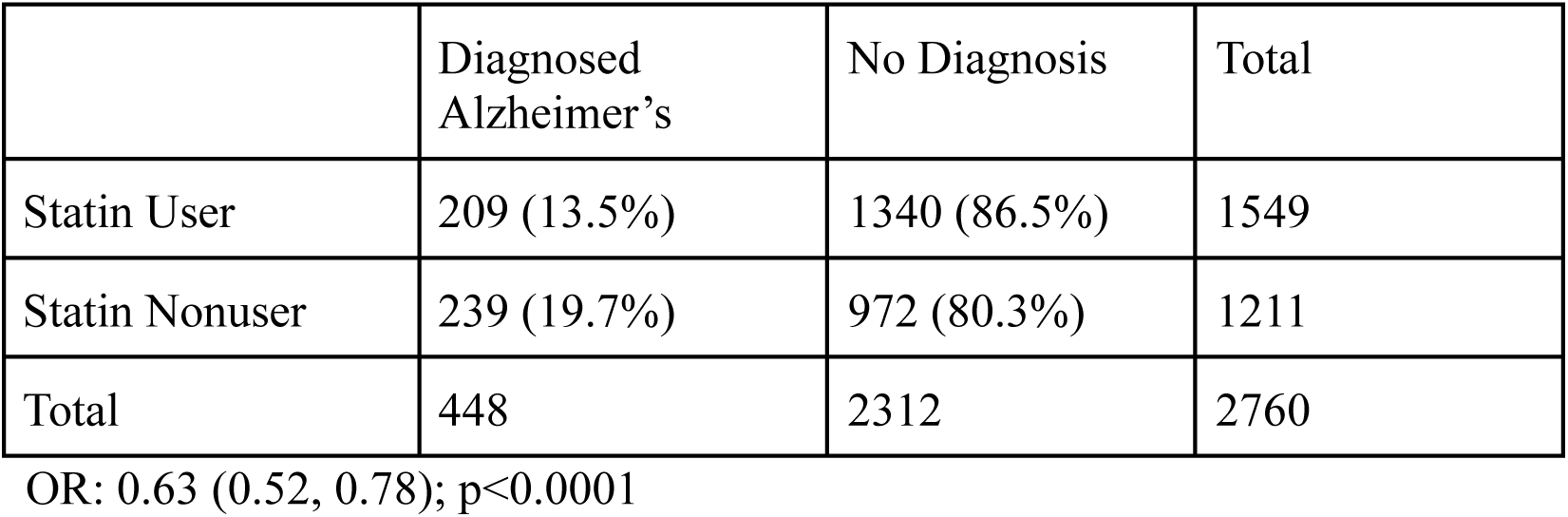
Diagnosis of Alzheimer’s disease among all statin users versus nonusers.

**Table 3:** Diagnosis of Other Dementias among all statin users versus nonusers.

|  | Diagnosed Dementia | No Diagnosis | Total |
| --- | --- | --- | --- |
| Statin User | 349 (20.7%) | 1340 (79.3%) | 1689 |
| Statin Nonuser | 359 (27.0%) | 972 (73.0%) | 1331 |
| Total | 708 | 2312 | 3020 |

OR: 0.71 (0.60, 0.83); $p < 0.0001$

As part of a sub-analysis comparing ADRD risk reductions associated with individual statins, **Table 4** features crosstabulations comparing ADRD diagnosis among users of atorvastatin, simvastatin, pravastatin, and rosuvastatin with ADRD diagnosis among all statin nonusers. For this sub-analysis, respondents were deemed users of whichever statin they were dispensed over 55% of the time. As a result, 5 statin users were excluded from the sub-analysis, each having had either two (n=4) or three (n=1) different statin drugs dispensed to them equally. Each of the four statin drugs investigated were correlated with significantly reduced ADRD risk, with atorvastatin, simvastatin, pravastatin, and rosuvastatin associated with 22.2% (OR 0.78, 95% CI: 0.66-0.99; p=0.0040), 37.5% (OR 0.63, 95% CI: 0.50-0.79; p<0.0001), 41.8% (OR 0.58, 95% CI: 0.42-0.80; p=0.0007), and 48.6% (OR 0.51, 95% CI: 0.37-0.72; p<0.0001) reductions in odds of ADRD diagnosis, respectively.

**Table 4:** Diagnosis of ADRD among atorvastatin, simvastatin, pravastatin, and rosuvastatin users versus all statin nonusers.

|  | Diagnosed ADRD | No Diagnosis | Total | OR (relative to nonusers) with p-value |
| --- | --- | --- | --- | --- |
| Atorvastatin User | 306 (32.4%) | 639 (67.6%) | 945 | 0.78 (0.66, 0.99);<br>p=0.0040 |
| Simvastatin User | 120 (27.7%) | 312 (72.3%) | 432 | 0.63 (0.50, 0.79);<br>p<0.0001 |
| Pravastatin User | 58 (26.4%) | 162 (73.6%) | 220 | 0.58 (0.42, 0.80);<br>p=0.0007 |
| Rosuvastatin User | 49 (24.0%) | 155 (76.0%) | 204 | 0.51 (0.37, 0.72);<br>p<0.0001 |
| Statin Nonuser | 598 (38.1%) | 972 (61.9%) | 1570 |  |
| Total | 1131 | 2240 | 3371 |  |

**Table 5** displays odds ratios and corresponding p-values of ADRD diagnosis among users of one particular statin drug (simvastatin, pravastatin, or rosuvastatin) relative to diagnosis among users of a different statin drug (atorvastatin, simvastatin, or pravastatin). Rosuvastatin was associated with significantly reduced ADRD risk relative to atorvastatin, with exposure to the former reducing the odds of ADRD diagnosis by 34.0% (OR 0.66, 95% CI: 0.47-0.94; p=0.0193) relative to exposure to the latter.

**Table 5:** Treatment effect comparison matrix - odds of ADRD diagnosis among users of treatment versus control statin.

| Control | Treatment |  |  |
| --- | --- | --- | --- |
|  | Simvastatin | Pravastatin | Rosuvastatin |
| Atorvastatin | 0.80 (p=0.0901) | 0.75 (p=0.0900) | **0.66 (p=0.0193)** |
| Simvastatin |  | 0.93 (p=0.7804) | 0.82 (p=0.3373) |
| Pravastatin |  |  | 0.88 (p=0.6546) |

Finally, **Table 6** displays a crosstabulation comparing ADRD diagnosis among users of the two hydrophilic statins, pravastatin and rosuvastatin, with ADRD diagnosis among users of lipophilic statins (atorvastatin and simvastatin, as well as fluvastatin, lovastatin, and pitavastatin). Overall, hydrophilic statins were associated with significantly reduced ADRD risk relative to lipophilic statins, with exposure to hydrophilic statins reducing the odds of ADRD diagnosis by 23.1% (OR 0.77, 95% CI: 0.60-0.98; p=0.0394) relative to exposure to lipophilic statins.

**Table 6:** Diagnosis of ADRD among users of hydrophilic statins versus users of lipophilic statins.

|  | Diagnosed ADRD | No Diagnosis | Total |
| --- | --- | --- | --- |
| Hydrophilic Statin<br>(Pravastatin,<br>Rosuvastatin) | 107 (25.2%) | 317 (74.8%) | 424 |
| Lipophilic Statin<br>(Atorvastatin,<br>Simvastatin, Fluvastatin,<br>Lovastatin, Pitavastatin) | 448 (30.5%) | 1021 (69.5%) | 1469 |
| Total | 555 | 1338 | 1893 |

OR: 0.77 (0.60, 0.98); p=0.0394

## DISCUSSION

Statin utilization was associated with significantly reduced risk of ADRD diagnosis among the studied cohort of Medicare beneficiaries, regardless of which statin drug (atorvastatin, simvastatin, pravastatin, or rosuvastatin) was utilized. This finding was consistent with Barthold et al.’s previous conclusion that atorvastatin, simvastatin, pravastatin, and rosuvastatin, all significantly reduced ADRD risk when taken in combination with angiotensin receptor blockers (ARBs) for hypertension.^23^ When comparing the degrees to which each of the four investigated statins were associated with reduced ADRD risk, we found that rosuvastatin was associated with significantly reduced risk relative to atorvastatin. This result also coincided with Barthold et al.’s earlier finding that rosuvastatin was significantly more effective at reducing ADRD risk than atorvastatin when both were taken in combination with angiotensin-converting enzyme inhibitors (ACEIs) for hypertension.^23^ Lastly, when comparing the efficacies of hydrophilic and lipophilic statins in reducing ADRD risk, hydrophilic statins (pravastatin and rosuvastatin) were observed to correlate with significantly reduced ADRD risk relative to lipophilic statins (atorvastatin, simvastatin, fluvastatin, lovastatin, pitavastatin). This finding was again congruent with Barthold et al.’s conclusion that pravastatin (when used in combination with ARBs) and rosuvastatin (when used in combination with ACEIs) were significantly more effective at reducing ADRD risk than both atorvastatin and simvastatin when used in combination with each of the respective antihypertensives, as well as Sinyavskaya et al.’s conclusion that lipophilic statins were associated with higher risk of Alzheimer’s disease compared to hydrophilic statins.^23,24^

During study conceptualization, we hypothesized that the degree to which a particular statin drug would correlate to reduced ADRD risk would be predicated upon its relative effectiveness at treating dyslipidemia. In 2010, Weng et al.’s systematic review and meta-analysis of 75 studies on the therapeutic equivalence of statins found that rosuvastatin and atorvastatin were significantly more effective at treating dyslipidemia than other statin drugs, each reducing LDL-C by more than 40%.^31^ Zhang et al.’s 2020 systematic review and network meta-analysis of 50 randomized controlled trials confirmed this finding, ranking rosuvastatin, atorvastatin, and pitavastatin as the first, second, and third most effective statins at lowering LDL-C.^32^ In contrast, while both this analysis and Barthold et al.’s found rosuvastatin to correlate to significantly reduced ADRD risk relative to other statin drugs, both analyses also found exposure to pravastatin to correlate to greater ADRD risk reductions than exposure to atorvastatin.^23^ As rosuvastatin and pravastatin are both hydrophilic, while atorvastatin is lipophilic, this incongruity leads us to conclude that hydrophilicity in statin medications plays a far more important role in ADRD prevention than in controlling dyslipidemia.

A major limitation to this analysis, of course, was that, given the cross-sectional nature of the utilized MCBS data, temporality could not be established between initiation of statin therapy and diagnosis of ADRD. Although the MCBS has been conducted annually since 1991, its rotating panel design does not allow for longitudinal analysis beyond four years.^33^ Additionally, the epidemiologic research methods used in this analysis do not aid in facilitating a better understanding of why the hydrophilic natures of certain statin drugs play an instrumental role in reducing ADRD risk; this phenomenon continues to evade researchers in the space, with Jamshidnejad-Tosaramandani et al. writing as recently as 2022 that “the difficulty in explaining the influence of statin lipophilicity on cognition can be ascribed to the diverse effects of statins on different types of dementia based on their lipophilicity.”^34^

## CONCLUSIONS

In conclusion, our findings coincide with others’ and suggest that: a) statins as an overall drug class correlate to significantly reduced ADRD risk, and b) hydrophilic statins (pravastatin and rosuvastatin) correlate to significantly greater ADRD risk reduction relative to lipophilic statins. However, there remains a need for future longitudinal studies, either prospective or retrospective, to establish temporality between statin therapy initiation and ADRD diagnosis to deem statins an effective source of prophylaxis against ADRD. In addition, given the present lack of understanding behind how hydrophilicity impacts the prophylactic efficacies of certain statins, and, more broadly, of the mechanism(s) by which statins interfere with the Aβ production pathway, further basic scientific research aimed at studying the biochemical and pharmacological connections between statins and ADRD deterrence is also warranted. Regardless, physicians intending to initiate a patient on statin therapy for uncontrolled dyslipidemia should consider prescribing rosuvastatin in lieu of atorvastatin for the additional prophylactic benefits against ADRD offered by the former, while patients apprehensive about initiating statin therapy should weigh into their decision making the added protection against Alzheimer’s disease and other forms of dementia that any prescribed statin should provide.

## AUTHOR CONTRIBUTIONS

MC, ZMK, and AA were all responsible for study design and data interpretation. MC conducted the analysis and wrote the first draft of the manuscript. All authors critically commented on the manuscript and approved the final version.

## DATA SHARING STATEMENT

Complete annual MCBS data sets from 2015 onward, excluding specific direct identifiers as defined by the HIPAA Privacy Rule, can be requested by researchers through filing a limited data set data use agreement (DUA) with CMS. For more information visit https://www.cms.gov/data-research/files-for-order/limited-data-set-lds-files/medicare-current-beneficiary-survey-mcbs.

## Data Availability

Complete annual MCBS data sets from 2015 onward, excluding specific direct identifiers as defined by the HIPAA Privacy Rule, can be requested by researchers through filing a limited data set data use agreement (DUA) with CMS.

https://www.cms.gov/data-research/files-for-order/limited-data-set-lds-files/medicare-current-beneficiary-survey-mcbs

## APPENDIX

### I.#Identification of ADRD Diagnosis Status

Beneficiary ADRD diagnosis statuses were identified through variables OCALZMER and OCDEMENT on the self-reported Health Status and Functioning Questionnaire, as published in MCBS’s CHRNCOND module, and variables I4200 and I4800 on the Health Status section of the MCBS Facility Instrument, as published in the MDS3 module.

### II.#Demographic & Matching

Sex, race (and, if possible, ethnicity), age stratum, and income level were used to match cases and controls. For context, the MCBS Demographics and Income Questionnaire lists several ethnicities to follow up with if a beneficiary responds as identifying as Asian, Hispanic, or Native Hawaiian or Pacific Islander. MCBS divides age into the following strata: 0-44, 45-64, 65-69, 70-74, 75-79, 80-84, and 85+.

While as of 2020, annual income has been stratified on the MCBS in $5,000 increments from $0 to $29,999, $10,000 increments from $30,000 to $59,999, and $20,000 increments from $60,000 to $139,999 (before terminating at $140,000+), income stratification on the 2019 survey terminated at $50,000+. In order to aggregate 2019 and 2020 demographic data for the purposes of this analysis, 2019 income level data was restratified into the updated income brackets using total annual income reported by responding beneficiaries.

Table A demographically breaks down the included cases by diagnosis type, sources of diagnosis data, and key characteristics (excluding ethnicity) used to match the cases with controls at 1:2 ratio, as well as education and hypertension prevalence data. 38.8% of the included cases reported diagnosis with Alzheimer’s disease, while the remaining 61.2% reported diagnosis with other forms of dementia. Sources of disease status data were split almost evenly among cases, with 56.5% of the diagnosis information having been self-reported via the MCBS Health Status and Functioning Questionnaire, and the remaining 43.5% having been reported via the MCBS Facility Instrument by representatives of long-term care facilities on behalf of their resident cases. Females comprised 65.3% of all selected cases, resulting in a slightly higher ratio of females-to-males than observed in the overall sample of MCBS respondents with available prescription utilization data (55.8% female). The included cases were also proportionally older than the overall sample of MCBS respondents, with 55.0% of all cases aged 85 years or older, compared to just 20.2% of all respondents with available prescription data. White respondents represented 75.2% of all included cases while Hispanic and Black respondents represented 12.0% and 9.7%, respectively. 56.0% of all included cases reported annual incomes ranging between $5,000 and $24,999, and 53.6% reported high school (including vocational/technical school) graduation or less as their highest attained level of education. Hypertension was prevalent in at least 69.6% of all included cases.

**Table A:** Demographic characteristics of respondents living with ADRD.

| Characteristic | Level | Count (n = 1156) |
| --- | --- | --- |
| Diagnosis Type | Alzheimer's | 448 (38.8%) |
|  | Other Dementia | 708 (61.2%) |
| Reporting Method | Self-Reported | 653 (56.5%) |
|  | Facility Survey | 503 (43.5%) |
| Sex | Male | 401 (34.7%) |
|  | Female | 755 (65.3%) |
| Race | White | 869 (75.2%) |
|  | Black | 112 (9.7%) |
|  | Hispanic | 139 (12.0%) |
|  | Asian/Pacific Islander | 26 (2.2%) |
|  | Native American | 10 (0.9%) |
| Age | 45-64 | 38 (3.3%) |
|  | 65-69 | 39 (3.4%) |
|  | 70-74 | 78 (6.7%) |
|  | 75-79 | 138 (11.9%) |
|  | 80-84 | 227 (19.6%) |
|  | 85+ | 636 (55.0%) |
| Income | \$0-\$4,999 | 25 (2.2%) |
| | \$5,000-\$9,999 | 165 (14.3%) |
| | \$10,000-\$14,999 | 235 (20.3%) |
| | \$15,000-\$19,999 | 153 (13.2%) |
| | \$20,000-\$24,999 | 100 (8.7%) |
| | \$25,000-\$29,999 | 78 (6.7%) |
| | \$30,000-\$39,999 | 102 (8.8%) |
| | \$40,000-\$49,999 | 68 (5.9%) |
| | \$50,000-\$59,999 | 47 (4.1%) |
| | \$60,000-\$79,999 | 73 (6.3%) |
| | \$80,000-\$99,999 | 43 (3.7%) |
| | \$100,000-\$119,999 | 21 (1.8%) |
| | \$120,000-\$139,999 | 11 (1.0%) |
| | \$140,000+ | 35 (3.0%) |
| Highest Attained Education | None | 20 (1.7%) |
|  | K-8 | 136 (11.8%) |
|  | Some High School | 124 (10.7%) |
|  | High School Graduate | 278 (24.0%) |
|  | Vocational/Technical School | 62 (5.4%) |
|  | Some College | 109 (9.4%) |
|  | Two-Year College Graduate | 47 (4.1%) |
|  | Four-Year College Graduate | 79 (6.8%) |
|  | Graduate School | 72 (6.2%) |
|  | Don't Know | 228 (19.7%) |
|  | No Answer | 1 (0.1%) |
| Hypertension | Yes | 805 (69.6%) |
|  | No | 262 (22.7%) |
|  | Don't Know | 88 (7.6%) |
|  | Refused | 1 (0.1%) |

Table B demographically breaks down the 2,312 controls included in the analysis by sex, race, age, income, attained education, and hypertension prevalence. Each of the selected controls matched their corresponding case by both race and sex. 54.2% of all selected controls were at least 85 years old, while 56.3% of controls reported annual incomes ranging between $5,000 and $24,999. Chi squared tests for goodness of fit indicated no significant differences between cases and selected controls in terms of age (χ^2^ = 1.621; p=0.8987), income (χ^2^ = 2.002; p=0.9998); ethnicity (χ^2^ = 4.176; p=0.9645), or hypertension (χ^2^ = 4.350; p=0.2261), prevalent in at least 69.7% of all selected controls, but did indicate significant differences in terms of highest attained education levels (χ^2^ = 388.115; p<0.0001). This observation, however, was primarily attributable to an abnormally high proportion of “Don’t Know” answers for highest attained education level for ADRD cases whose responses were provided by long-term care facility representatives (n=215 out of 228 total “Don’t Know” answers among ADRD cases).

**Table B:** Demographic characteristics of selected controls.

| Characteristic | Level | Count (n = 2312) |
| --- | --- | --- |
| Sex | Male | 802 (34.7%) |
|  | Female | 1510 (65.3%) |
| Race | White | 1738 (75.2%) |
|  | Black | 224 (9.7%) |
|  | Hispanic | 278 (12.0%) |
|  | Asian/Pacific Islander | 52 (2.2%) |
|  | Native American | 20 (0.9%) |
| Age | 45-64 | 70 (3.1%) |
|  | 65-69 | 82 (3.5%) |
|  | 70-74 | 159 (6.9%) |
|  | 75-79 | 288 (12.5%) |
|  | 80-84 | 460 (19.9%) |
|  | 85+ | 1253 (54.2%) |
| Income | \$0-\$4,999 | 49 (2.1%) |
| | \$5,000-\$9,999 | 318 (13.8%) |
| | \$10,000-\$14,999 | 469 (20.3%) |
| | \$15,000-\$19,999 | 307 (13.3%) |
| | \$20,000-\$24,999 | 205 (8.9%) |
| | \$25,000-\$29,999 | 157 (6.8%) |
| | \$30,000-\$39,999 | 207 (9.0%) |
| | \$40,000-\$49,999 | 133 (5.8%) |
| | \$50,000-\$59,999 | 91 (3.9%) |
| | \$60,000-\$79,999 | 153 (6.6%) |
| | \$80,000-\$99,999 | 93 (4.0%) |
| | \$100,000-\$119,999 | 41 (1.8%) |
| | \$120,000-\$139,999 | 23 (1.0%) |
| | \$140,000+ | 66 (2.9%) |
| Highest Attained Education | None | 35 (1.5%) |
|  | K-8 | 252 (21.8%) |
|  | Some High School | 302 (26.1%) |
|  | High School Graduate | 637 (55.1%) |
|  | Vocational/Technical School | 155 (13.4%) |
|  | Some College | 294 (25.4%) |
|  | Two-Year College Graduate | 98 (8.5%) |
|  | Four-Year College Graduate | 238 (20.6%) |
|  | Graduate School | 194 (16.8%) |
|  | Don't Know | 105 (9.1%) |
|  | No Answer | 2 (0.1%) |
| Hypertension | Yes | 1611 (69.7%) |
|  | No | 500 (21.6%) |
|  | Don't Know | 198 (8.6%) |
|  | Refused | 3 (0.1%) |

## Notes

### Competing Interest Statement

The authors have declared no competing interest.

### Funding Statement

No research funding was associated with this study.

## REFERENCES

1. Knopman DS, Amieva H, Petersen RC, et al. Alzheimer disease. Nat Rev Dis Primers. 2021;7(1):33.

2. Breijyeh Z, Karaman R. Comprehensive Review on Alzheimer’s Disease: Causes and Treatment. Molecules. 2020;25(24):5789.

3. Tackenberg C, Kulic L, Nitsch RM. Familial Alzheimer’s disease mutations at position 22 of the amyloid β-peptide sequence differentially affect synaptic loss, tau phosphorylation and neuronal cell death in an ex vivo system. PLoS One. 2020;15(9):e0239584.

4. Wang H, Kulas AJ, Wang C, Holtzman DM, Ferris HA, Hansen SB. Regulation of beta-amyloid production in neurons by astrocyte-derived cholesterol. PNAS. 2020;118(33):e2102191118.

5. Singh-Manoux A, Gimeno D, Kivimaki M, Brunner E, Marmot MG. Low HDL cholesterol is a risk factor for deficit and decline in memory in midlife: the Whitehall II study. Arterioscler Thromb Vasc Biol. 2008;28(8):1556–62.

6. Moreta MP, Burgos-Alonso N, Torrecilla M, Marco-Contelles J, Bruzos-Cidón C. Efficacy of Acetylcholinesterase Inhibitors on Cognitive Function in Alzheimer’s Disease. Review of Reviews. Biomedicines. 2021;9(11):1689.

7. Food and Drug Administration. Advancing Health Through Innovation: New Drug Therapy Approvals 2021. fda.gov. Available at: https://www.fda.gov/media/155227/download. Updated May 13, 2022. Accessed April 13, 2023.

8. Abyadeh M, Gupta V, Gupta V, et al. Comparative Analysis of Aducanumab, Zagotenemab and Pioglitazone as Targeted Treatment Strategies for Alzheimer’s Disease. Aging Dis. 2021;12(8):1964–1976.

9. Institute for Clinical and Economic Review. ICER Issues Statement on the FDA’s Approval of Aducanumab for Alzheimer’s Disease. icer.org. Available at: https://icer.org/news-insights/press-releases/icer-issues-statement-on-the-fdas-approval-of-aducanumab-for-alzheimers-disease/. Updated June 7, 2021. Accessed April 13, 2023.

10. Institute for Clinical and Economic Review. ICER Publishes Evidence Report on Lecanemab for Alzheimer’s Disease. icer.org. Available at: https://icer.org/news-insights/press-releases/icer-publishes-evidence-report-on-lecanemab-for-alzheimers-disease/. Updated March 1, 2023. Accessed April 13, 2023.

11. Eli Lilly and Company. U.S. Food and Drug Administration Issues Complete Response Letter for Accelerated Approval of Donanemab. investors.lilly.com. Available at: https://investor.lilly.com/news-releases/news-release-details/us-food-and-drug-administration-issues-complete-response-0. Updated January 19, 2023. Accessed April 13, 2023.

12. Lilly’s Kisunla™ (donanemab-azbt) Approved by the FDA for the Treatment of Early Symptomatic Alzheimer’s Disease. investors.lilly.com. Available at: https://investor.lilly.com/news-releases/news-release-details/lillys-kisunlatm-donanemab-azbt-approved-fda-treatment-early. Updated July 2, 2024. Accessed July 4, 2024.

13. Biogen Inc. Biogen Announces Reduced Price for ADUHELM® to Improve Access for Patients with Early Alzheimer’s Disease. investors.biogen.com. Available at: https://investors.biogen.com/news-releases/news-release-details/biogen-announces-reduced-price-aduhelmr-improve-access-patients. Updated December 20, 2021. Accessed April 14, 2023.

14. Eisai Co., Ltd. Eisai’s Approach to U.S. Pricing for LEQEMBI™ (Lecanemab), a Treatment for Early Alzheimer’s Disease, Sets Forth Our Concept of “Societal Value of Medicine” in Relation to “Price of Medicine.” eisai.com. Available at: https://www.eisai.com/news/2023/news202302.html. Updated January 7, 2023. Accessed April 14, 2023.

15. Centers for Medicare & Medicaid Services. CMS Finalizes Medicare Coverage Policy for Monoclonal Antibodies Directed Against Amyloid for the Treatment of Alzheimer’s Disease. cms.gov. Available at: https://www.cms.gov/newsroom/press-releases/cms-finalizes-medicare-coverage-policy-monoclonal-antibodies-directed-against-amyloid-treatment. Updated April 7, 2022. Accessed April 14, 2023.

16. Taylor F, Huffman MD, Macedo AF, et al. Statins for the primary prevention of cardiovascular disease. Cochrane Database Syst Rev. 2013;2013(1):CD004816.

17. Product Name: Atorvastatin Calcium. RED BOOK Online. IBM Micromedex [database online]. Truven Health Analytics/IBM Watson Health; 2023. Accessed January 12, 2024. https://www.micromedexsolutions.com.

18. Product Name: Pravastatin Sodium. RED BOOK Online. IBM Micromedex [database online]. Truven Health Analytics/IBM Watson Health; 2023. Accessed January 12, 2024. https://www.micromedexsolutions.com.

19. Product Name: Rosuvastatin Calcium. RED BOOK Online. IBM Micromedex [database online]. Truven Health Analytics/IBM Watson Health; 2023. Accessed January 12, 2024. https://www.micromedexsolutions.com.

20. Product Name: Simvastatin. RED BOOK Online. IBM Micromedex [database online]. Truven Health Analytics/IBM Watson Health; 2023. Accessed January 12, 2024. https://www.micromedexsolutions.com.

21. Prospective Studies Collaboration, Lewington S, Whitlock G, et al. Blood cholesterol and vascular mortality by age, sex, and blood pressure: a meta-analysis of individual data from 61 prospective studies with 55,000 Lancet. 2008 Jul 26;372(9635):292.

22. Torrandell-Haro G, Branigan GL, Vitali F, Geifman N, Zissimopoulos JM, Brinton RD. Statin therapy and risk of Alzheimer’s and age-related neurodegenerative diseases. Alzheimers Dement (N Y). 2020;6(1):e12108.

23. Barthold D, Joyce G, Diaz Brinton R, Wharton W, Kehoe PG, Zissimopoulos J. Association of combination statin and antihypertensive therapy with reduced Alzheimer’s disease and related dementia risk. PLoS One. 2020 Mar 4;15(3):e0229541

24. Sinyavskaya L, Gauthier S, Renoux C, Dell’Aniello S, Suissa S, Brassard P. Comparative effect of statins on the risk of incident Alzheimer disease. Neurology. 2018 Jan 16;90(3):e179–e187.

25. Jonk Y, O’Connor H, Schult T, Cutting A, Feldman R, Ripley DC, Dowd B. Using the Medicare Current Beneficiary Survey to conduct research on Medicare-eligible veterans. J Rehabil Res Dev. 2010;47(8):797–813.

26. Centers for Medicare and Medicaid Services. Medicare Current Beneficiary Survey (MCBS).cms.gov. Available at: https://www.cms.gov/Research-Statistics-Data-andSystems/Research/MCBS. Updated February 8, 2023. Accessed April 19, 2023.

27. Centers for Medicare and Medicaid Services. MCBS Tutorials: Introduction for New Users. cms.gov. Available at: https://www.cms.gov/Research-Statistics-Data-andSystems/Research/MCBS/Downloads/MCBS_Tutorial.pdf. Updated November 23, 2021. Accessed April 19, 2023.

28. Centers for Medicare and Medicaid Services. 2020 MCBS Survey File. cms.gov. Available at: https://www.cms.gov/research-statistics-data-and-systemsresearchmcbscodebooks/2020-mcbs-survey-file. Updated June 29, 2022. Accessed April 20, 2023.

29. Centers for Medicare and Medicaid Services. Minimum Data Set (MDS) 3.0 for Nursing Homes and Swing Bed Providers. cms.gov. Available at: https://www.cms.gov/medicare/quality-initiatives-patient-assessment-instruments/nursinghomequalityinits/nhqimds30. Updated March 29, 2023. Accessed April 20, 2023.

30. Centers for Medicare and Medicaid Services. 2019 MCBS Survey File. cms.gov. Available at: https://www.cms.gov/research-statistics-data-and-systemsresearchmcbscodebooks/2019-mcbs-survey-file. Updated January 23, 2023. Accessed June 27, 2023.

31. Weng TC, Yang YH, Lin SJ, Tai SH. A systematic review and meta-analysis on the therapeutic equivalence of statins. J Clin Pharm Ther. 2010 Apr;35(2):139–51.

32. Zhang X, Xing L, Jia X, Pang X, Xiang Q, Zhao X, Ma L, Liu Z, Hu K, Wang Z, Cui Y. Comparative Lipid-Lowering/Increasing Efficacy of 7 Statins in Patients with Dyslipidemia, Cardiovascular Diseases, or Diabetes Mellitus: Systematic Review and Network Meta-Analyses of 50 Randomized Controlled Trials. Cardiovasc Ther. 2020 Apr 23;2020:3987065.

33. Centers for Medicare and Medicaid Services. MCBS Advanced Tutorial on Longitudinal Analysis Using MCBS Data. cms.gov. Available at: https://www.cms.gov/files/document/mcbs-advanced-tutorial-longitudinal-analysi <u>s.pdf</u>.

34. Jamshidnejad-Tosaramandani T, Kashanian S, Al-Sabri MH, Kročianová D, Clemensson LE, Gentreau M, Schiöth HB. Statins and cognition: Modifying factors and possible underlying mechanisms. Front Aging Neurosci. 2022 Aug 15;14:968039.

